# Real-world Evidence for Preventive Effects of Statins on Cancer Incidence: A Trans-Atlantic Analysis

**DOI:** 10.1101/2021.07.20.21260891

**Authors:** Bjoern-O Gohlke, Fabian Zincke, Andreas Eckert, Dennis Kobelt, Saskia Preissner, Juliane Maria Liebeskind, Nikolas Gunkel, Kerstin Putzker, Joe Lewis, Sally Preissner, Benedikt Kortüm, Wolfgang Walther, Cameron Mura, Philip E. Bourne, Ulrike Stein, Robert Preissner

## Abstract

**Background:** Numerous clinical trials have considered the potential linkages between statins and cancer. Despite some evidence for reduced mortality associated with statin use, the results thus far have been somewhat inconclusive and not easily comparable, thus hampering the emergence of a consensus. We suspect that this uncertainty would be reduced, and greater clarity achieved (e.g. regarding clinical best practices and standards-of-care), were we to have a reliable, causal biomarker that could help identify those individual patients who might benefit from statin use during cancer treatment.

**Methods and Findings:** In the joint experimental and statistical analysis reported here, we assessed the inhibitory potential of various statins on the expression of a tumor enhancer known as MACC1, taking into account the molecular functions of this key metastasis-associated protein. To assess any effects of statins in cancer prevention (observationally), we also performed a retrospective, two-center, nested case-control study, focusing on medical centers in Berlin, Germany and Virginia, USA. Among nearly a half-million patient visits, over a decade-long period, cancer patients were identified and analyzed in comparison to patients without cancer diagnoses. Odds ratios (OR) and hazard ratios (HR) for cancer were computed for patients with and without statin intake, accounting for potential confounders. Finally, we also extended these analyses of our trans-Atlantic cohort by utilizing real-world data from 132,072 cancer patients with statins available on the TriNetX platform.

Experimental work revealed that statins inhibit MACC1 mRNA levels and protein expression, resulting in reduced MACC1-induced phenotypic functions, such as motility and proliferation. Moreover, we found that statins restrict colorectal cancer (CRC) growth and metastasis in xenografted mice. The cohort data that we gathered at the German and U.S. centers enabled analysis of 53,113 cancer patients and matched controls. These were extracted, aggregated, and 1:1 matched (by age/gender) in order to build propensity-score matched sub-cohorts, to mitigate confounder bias. Based on this real-world evidence (RWE), we found that atorvastatin, fluvastatin, lovastatin, pravastatin, rosuvastatin and simvastatin were associated with a 50% reduced overall risk for developing cancer (OR 0.5, CI 0.48-0.51). The strongest association of reduced cancer risk was found for (i) liver cancer (OR 0.35, 0.29-0.43), (ii) secondary neoplasms of respiratory and digestive organs (OR 0.42, 0.34-0.45), and (iii) colorectal cancer (OR 0.44, 0.39-0.5). The effect of atorvastatin (OR 0.3, 0.28-0.32) exceeded other considered statins, even after exclusion of aspirin as the strongest confounder (OR 0.63, CI 0.57-0.7). Additionally, we note that those patients taking statins have a 38% decreased risk of death (HR 0.64, 0.48-0.86).

**Conclusions:** Our data, which offer evidence for cancer-preventative and anti-metastatic effects of statins, lead us to suggest that these medications should be considered in treating some types of cancers. In addition, MACC1 may serve as a potentially helpful biomarker for purposes of patient stratification (and personalized treatment). A more definitive test of these proposed ideas could come from prospective, randomized clinical trials.

## Introduction

Cancer is the second leading cause of death worldwide, claiming nearly ten million lives in 2020 according to the World Health Organization’s *International Agency for Cancer Research* [1]. The development of metastases is a particularly severe challenge in cancer therapy, given the aggressive nature of metastatic neoplasms and their associated pathophysiological processes, resulting in recalcitrance to treatment [2-4]; this difficulty underscores the need for alternative intervention strategies, such as chemoprevention, novel therapeutic targets, and new/repositioned drugs. We previously discovered that lovastatin restricts colorectal cancer (CRC) growth and metastasis, *in vitro* and *in vivo*, via the transcriptional inhibition of a tumor-promoting and metastasis-inducing gene known as “metastasis-associated in colon cancer 1” (MACC1) [5]. Lovastatin and other statins comprise a class of lipid– and cholesterol–lowering drugs that are effective in treating chronic cardiovascular conditions like hypercholesterolemia; these compounds act by inhibiting HMG-CoA reductase, thereby targeting the rate-limiting step of the biosynthetic mevalonate pathway [6-9]. In our present drug screening we identified a second statin compound, fluvastatin, as also being a transcriptional inhibitor of MACC1, prompting us to now assess the influence of all statin family members on MACC1-associated function *in vitro* and *in vivo*. Relative to our prior work [5], in the present study we have tested additional entities and functions *in vitro* as well as lower statin concentrations *in vivo*. To both broaden the scope of our study and gain a more thorough picture of possible statin ↔ cancer linkages, we used a real-world evidence (RWE) approach to examine potential associations between statin use and prevalence of various cancers among patients with positive diagnoses; this study design is necessarily observational, retrospective and cohort-based.

The current literature exhibits great variability as regards the reported effects of statins [7,10-12], and it is not straightforward to ascertain statistically significant results that clearly establish an effect. Such is the case largely because the significance of a set of results depends upon various confounding factors being recognized and accounted for (and, ideally, controlled for), such as small cohort sizes, co-medications, comorbidities, neglecting to distinguish different cancer types, confusion stemming from unknown or uncertain mechanisms of action, and even cohort-composition itself. For example, Mamtani et al. [13] examined the impact of *indication bias*—a type of bias wherein the indication itself (e.g., high cholesterol) may be causally(/mechanistically) associated with the outcome being studied (e.g., CRC or another cancer), thereby masking or otherwise muddying any inferences that can be drawn about the efficacy of the treatment/medication (e.g. statins). Confounders and biases are more difficult to handle in causal analyses of observational, retrospective studies (e.g. RWE), versus more traditional prospective, blinded, randomized clinical trials (RCT); an illuminating review of the RCT and RWE approaches was recently published by Eichler et al. [14].

Despite the shortcomings of the RWE approach, the ever-increasing availability of primary data on patients—particularly in well-structured, machine-readable forms, e.g. via standards-compliant electronic health records (EHR)—presents new opportunities for a more nuanced approach to drug discovery, as well as elucidation of the interrelationships between (i) a particular ailment, (ii) an individual patient’s history, comorbidities, etc., and (iii) the therapeutic(s) that might be most confidently indicated. Enabled by large data repositories, entire *populations* of patient that exhibit various underlying conditions can now be analyzed in RWE studies; doing so mitigates the pursuit of therapeutic approaches that are less likely to benefit a given patient, and would also enable better assessment of the significance of general conclusions drawn from large-scale retrospective studies [15,16].

In the work reported here, we have used a general RWE approach to test the hypothesis that statins have a cancer-preventive effect. Specifically, we conducted a RWE-based study of patients from the Charité–Universitätsmedizin Berlin and the University of Virginia (UVA) hospitals, both separately and as a single, combined cohort. To further broaden our analysis, we also examined a large, international cohort by using the TriNetX platform; this resource offers a federated health research network that provides access to EHR-based data (diagnoses, procedures, medications, laboratory values, genomic information, etc.), in a combined and de-identified manner (counts and aggregate statistical summaries). We present RWE for a preventive effect of statins based on 53,000 cancer patients from these two independent trans-Atlantic cohorts; reassuringly, the results from the TriNetX analysis, reflecting another 132,072 patients, are consistent with the single-site findings at Charité and UVA. At a molecular and mechanistic level, we provide experimental evidence that statins act, at least partly, by inhibiting transcription of the tumor-promoting and metastasis-inducing *MACC1* gene. We demonstrate this in vitro and in a model for CRC metastasis in xenografted mice.

## Methods

### Experimental Methods

High-throughput screens (HTS) of drug compounds were performed using HCT116 cells expressing the luciferase reporter gene, under the control of a *MACC1* promoter. Our library consisted of 4,241 compounds from Prestwick, NIH and Microsource. The best hits were validated by quantitative real-time polymerase chain reaction (qRT-PCR) and Western blot (WB) in cell lines of different cancer entities. To analyze cell proliferation, we used the IncuCyte® ZOOM System (Essen BioScience, Ann Arbor, Michigan). Colony forming abilities were analyzed with the ImageJ software (version 1.51j8, NIH, USA).

For *in vivo* assessment of our findings, we treated intra-splenically xenografted SCID-beige mice with daily doses of either solvent (negative control) or a statin. Specifically, the statins fluvastatin and atorvastatin were used in separate experiments, at a level of 13 mg/kg body weight. Note that this dosage level corresponds to a human equivalent dose of approximately 1 mg/kg body weight, which is a dose employed in blood lipid reduction therapy [17,18]. Tumor growth and metastasis formation were continuously monitored by bioluminescence imaging until the ethical end-point (cancer burden) was reached at 24 days. Animal experiments were conducted in compliance with the United Kingdom’s Coordinating Committee of Cancer Research (UKCCCR) guidelines, and approval was granted by the State Office of Health and Social Affairs (Landesamt für Gesundheit und Soziales, LaGeSo, Berlin, Germany) under the permit Reg0010/19.

### Study Population

The total population sampled in our study included patients from the Charité and UVA hospitals. During our initial data-processing steps, all diagnoses and discharge medications were extracted, merged, unified and transformed into a centralized resource that we developed in-house using an open-source, relational database management system (MySQL). Discharge medications—i.e., those which are actively prescribed at discharge/exit from a visit—were extracted from doctor letters at the Charité and from EHRs at UVA. A condition of ‘cancer’ was defined based on the ICD10 diagnosis codes C00 to D48, with exclusion of (i) codes D10 → D37 (which describe benign neoplasms) as well as (ii) newborns or those individuals older than 100 years [19,20]. This data-preparation process (Figure 1) yielded 277,980 distinct patients, of whom 53,113 were diagnosed with cancer (Table 1, Supp Figure 1). All U.S. patient records were anonymized prior to study cohort selection, in accordance with the Health Insurance Portability and Accountability Act (HIPAA) [21].

**Table 1:**
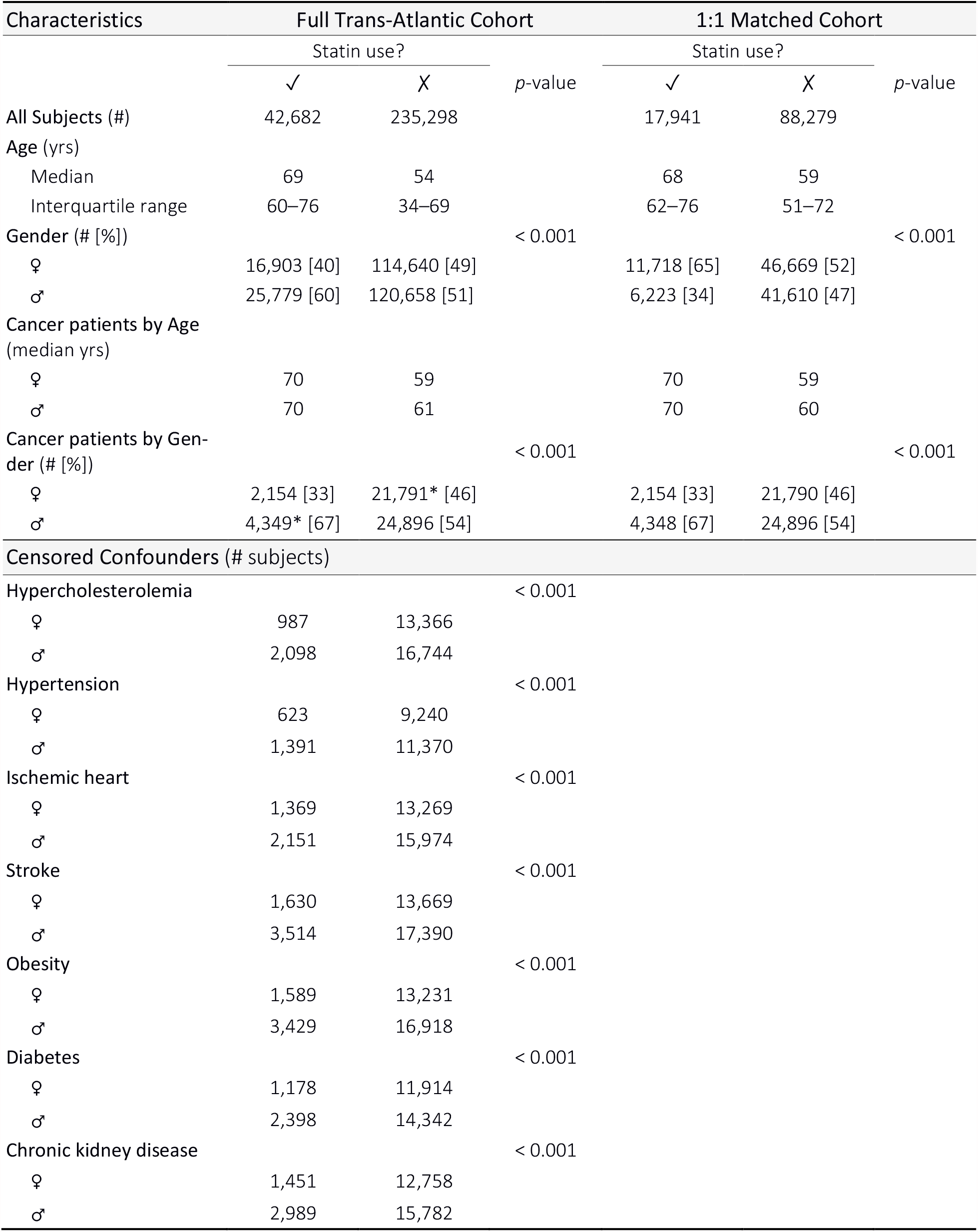
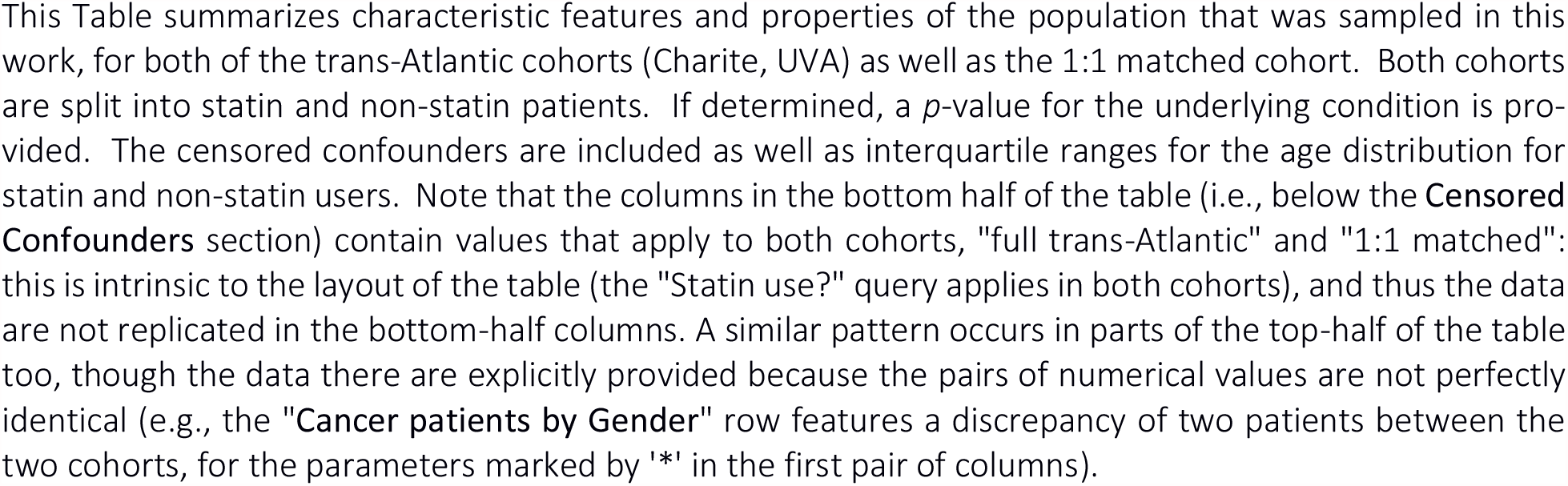
Statistical characteristics of the sampled population and study design.

**Figure 1:**
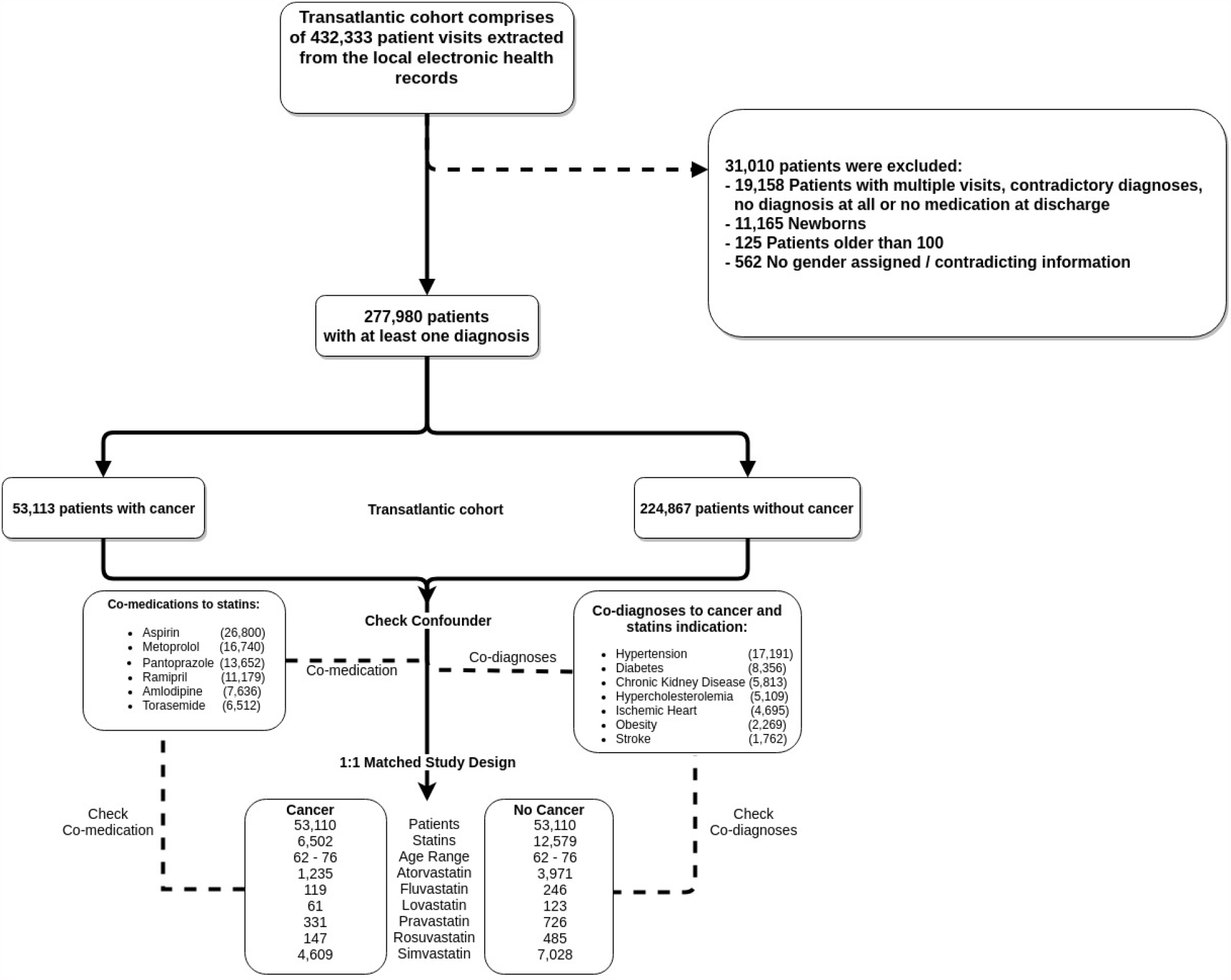
Data pre-processing and cohort design. Patient records from the repositories at the Charité and UVA Health System were merged into a transatlantic cohort. From the resulting 432,333 records, 31,010 were excluded (upper-right panel) due to incompleteness or contradictory information; this pre-processing yielded a final transatlantic cohort of 277,980 patients. Of these patients, 53,113 were diagnosed with cancer, where ‘cancer’ is defined based on ICD10 codes in the range of ‘C00’ to ‘D48’ (excluding codes ‘D10’ to ‘D37’, which describe benign neoplasms). Co-medications prescribed along with statins (lower-left), as well as co-diagnoses with cancer and statin indications (lower-right), were included in the confounder analysis and calculated for both the full transatlantic and 1:1 matched cohorts.

### Trial Design, Cohort Properties

We scrutinized the RWE for treatment of cancer with statins in an observational, large-scale, multi-center, retrospective cohort–based study. Charité and UVA datasets were separately analyzed, and were also considered as a combined trans-Atlantic cohort (Table 1). In terms of medications, in the early phase of this trans-Atlantic study patient statuses used in our analyses were those at discharge. To account for age and gender artifacts arising from patterns of statin prescription, patients with a cancer diagnosis were randomly matched 1:1 with non-cancer patients based on age and gender (right-half of Table 1). Cancer entities with large case counts were analyzed separately. The same methodology was also applied to different statins (individual compounds) in order to assess their chemopreventive potential (Figure 1).

### Confounder Effects

Various confounding factors were considered in this study. Potential biases stemming from familial history, and prescription or diagnosis habits of physicians, was minimized (‘averaged out’) by the trans-Atlantic design of this study (i.e., sampling over populations in Virginia and Germany). To mitigate the risk of describing cancer-preventive effects of statins as false-positives, the impact of drugs most commonly prescribed along with statins (i.e., co-medications) for each patient were examined. We also analyzed whether co-diagnoses based on ICD10 codes influence the conclusions reached in our study.

### Statistical Analyses

Odds ratios (OR) and risk ratios (RR) were calculated for the respective case and control groups [4,6]. To account for the study design, a 1:1 matched analysis was performed, taking into consideration the contingency table for different statins and cancer subtypes. The matched cohort was created by matching cancer patients by age and gender to non-cancer patients. To overcome possible bias by creating the control group (non-cancer patients), this procedure was performed 25 times, and ORs and RRs were considered as mean and median values.

In addition to the calculated ORs and RRs, we also performed a statistical survival analysis to investigate the effect of statins in cancer therapies. To assess the significance and validity of our results, we performed a log-rank test for the survival distribution of cancer patients taking statins and not taking statins. Specifically, we calculated hazard ratios (HR) by using a Cox-proportional hazard model.

All measurements are reported within 95% confidence intervals (CIs) and two-sided *p*-values, with a *p* < 0.05 considered statistically significant.

We analyzed the mRNA expression data statistics with a one-way analysis of variance (ANOVA) procedure, including Dunnett’s multiple comparison test; the functional assays were analyzed using two-way ANOVA including Tukey’s additivity test (both within a CI of 95%).

## Results

### Statins Inhibit MACC1 Expression and Function in Cell Culture and Metastasis Formation in Mice

When present at anomalously high levels, MACC1, an established prognostic and predictive biomarker, is known to aggravate tumor growth and induce metastasis formation in several solid tumor entities, including CRC, pancreatic and gastric cancers [22]. Thus, we evaluated the effects of different statins on MACC1, as a molecular target previously identified by our group [5,23]. We used HCT116 cells stably expressing the MACC1-promoter driven luciferase gene (Figure 2A) in a HTS to identify novel transcriptional inhibitors of MACC1. A stepwise process, including cytotoxicity, selectivity assessment (counter screen with HCT116/CMVp-Luc cells) and in vitro validation, confirmed fluvastatin as the most promising transcriptional inhibitor of MACC1 (Figure 2B). Our real-world data (described below) prompted us to assess the influence of additional statins on MACC1 expression. Fluvastatin as well as lovastatin, atorvastatin and simvastatin reduced the expression of MACC1 on mRNA and protein level in a concentration-dependent manner (Figure 2C). In a cross-entity approach, we confirmed the inhibitory effects of statins in pancreatic (BxPC3) and gastric (MKN45) cancer cells (Supp Figure 4).

**Figure 2:**
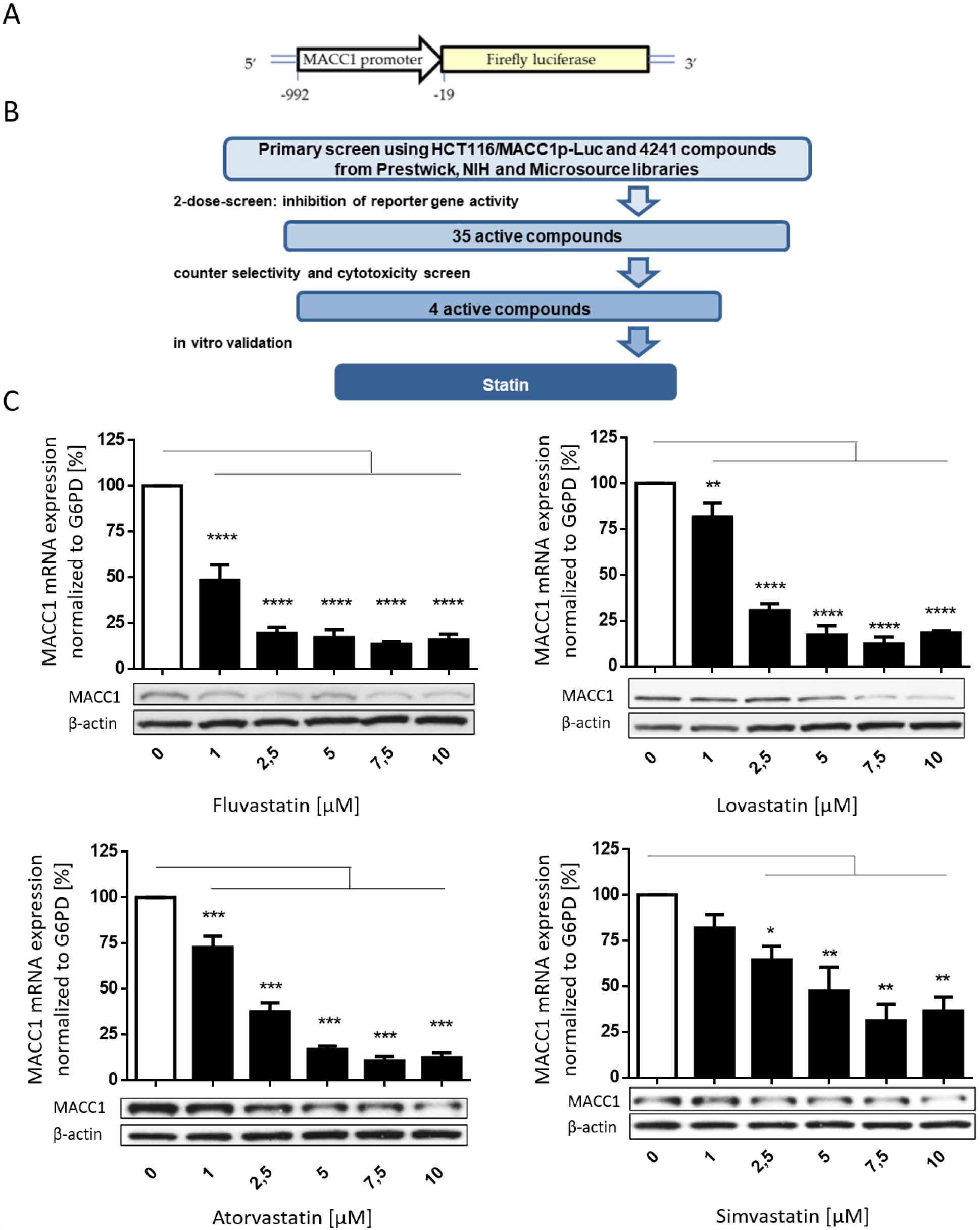
Statins reduce MACC1 expression. This schematic (A) illustrates the vector expressed in HCT116/ MACC1p-Luc cells that were employed in our HTS. The luciferase reporter gene is controlled by the MACC1 promoter (MACC1p). A tiered process revealed fluvastatin as transcriptional inhibitor of MACC1, with a total of 4,241 compounds tested. Initial 2-dose screening was followed by cytotoxicity and selectivity (with HCT116/CMVp-Luc cells) assessment as well as in vitro validation (B). Dose-dependent reduction of MACC1 mRNA and protein expression in HCT116 cells by different statins (fluvastatin, atorvastain, lovastatin, simvastatin) is shown in (C). MACC1 mRNA levels were normalized to G6PD mRNA expression and respective treatment controls (DMSO, indicated with white bars). Results for mRNA represent means ± standard error of the mean (SEM) of three independent experiments; for WB, one representative example of at least two independent experiments is shown. In the WB, β-actin or vinculin served as loading controls. Significant results were determined by one-way ANOVA and Dunnett’s multiple comparison test with a 95% CI (* = *p* < 0.05, ** = *p* < 0.01, *** = *p* < 0.001, **** = *p* < 0.0001).

We evaluated the potential of statins to inhibit MACC1-dependent proliferation and clonogenicity. Live-cell imaging showed that statin treatment remarkably reduces proliferation in control cells, whereas inducing MACC1 expression via a tetracycline-inducible promoter partly rescues this effect (Supp Figure 5A). Knocking-out MACC1 in HCT116 cells led to a severe decrease in proliferation, and this effect was not further reduced by statin treatment (Supp Figure 5B). To assess MACC1-dependent reproductive viability, we performed a clonogenic assay. Control cells (HCT116/GFP) demonstrated reduced colony formation upon statin treatment, but the overexpression of MACC1-GFP restored this effect (Supp Figure 5C-E).

For in vivo validation, we treated xenografted SCID-beige mice with either solvent, fluvastatin or atorvastatin (equating to a human dose of ca. 1 mg/kg body weight). Bioluminescence monitoring showed a drastic restriction of metastasis formation in statin-treated animals over the course of the experiment. Metastasis formation at day 24, compared to the control group, corresponded to a *p* < 0.0001 for fluvastatin and atorvastatin (Figure 3). Concomitantly, the levels of MACC1 transcript were found to be reduced in the livers of statin-treated mice (Figure 3C).

**Figure 3:**
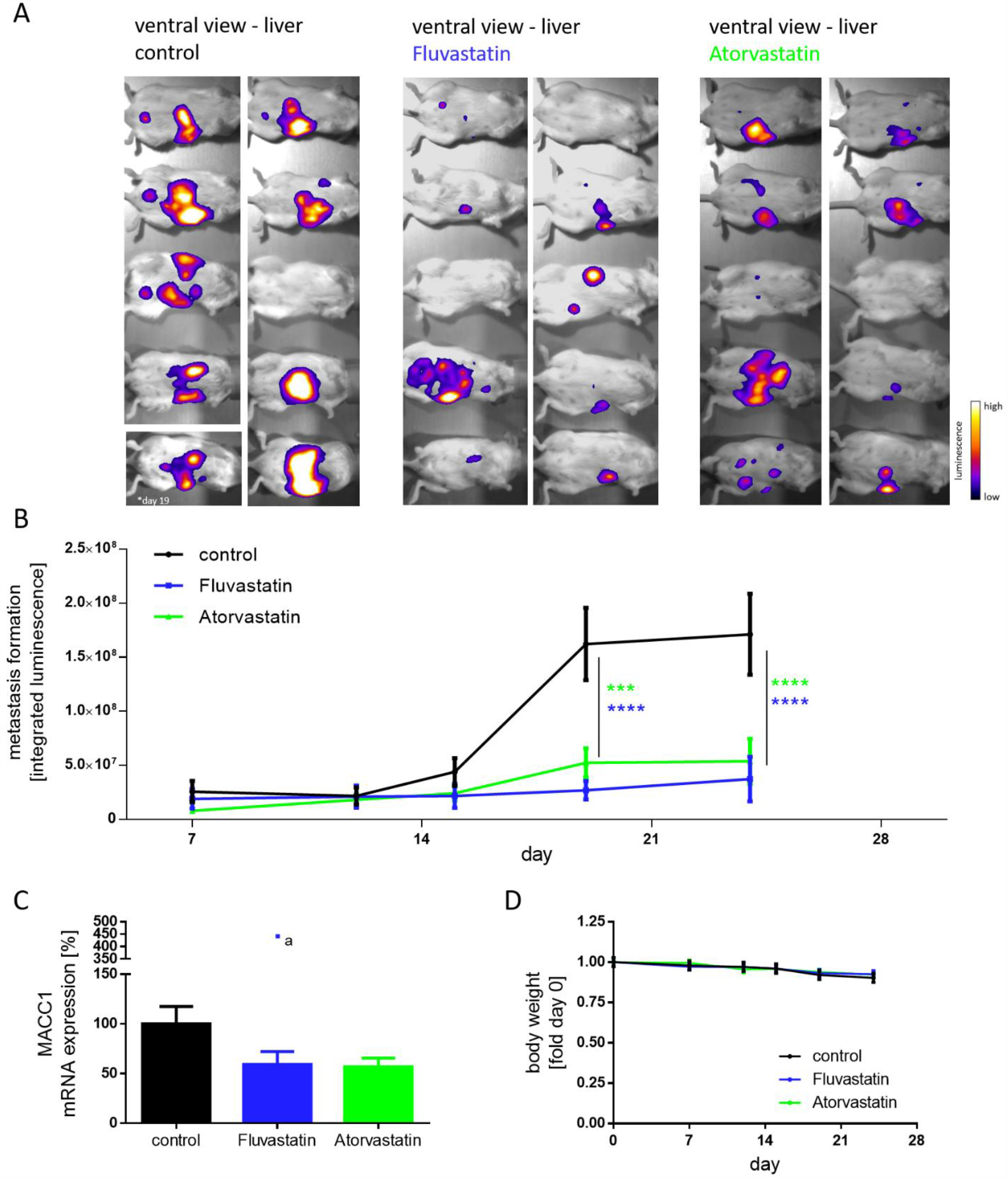
Statin treatment decreases tumor burden and metastasis formation in vivo. Intra-splenically xenografted (HCT116/CMVp-Luc cells) SCID-beige mice were treated either with solvent or daily doses of 13 mg/kg body weight fluvastatin or atorvastatin. Bioluminescent imaging of animals from a ventral view (A), at day 24 of statin treatment, showed significantly weaker signals, indicating restricted metastasis formation in the liver. Bioluminescent signals from all ten animals over the time course of the experiment were quantified (B). MACC1 transcripts in the liver were also found to be reduced in the fluvastatin and atorvastatin treated groups (C). Constant body weight of animals in each group indicates ethical conditions throughout the experiment (D). Results represent means ± SEM and significant results were determined by two-way ANOVA and Dunnet’s multiple comparison test with a 95% CI (* = *p* < 0.05, ** = *p* < 0.01, *** = *p* < 0.001, **** = *p* < 0.0001). One outlier, marked by an *a in the plot, was identified by the ROUT method (Q=1%) and Grubbs test (alpha=0.01); that datum was not included in calculating the mean MACC1 mRNA expression levels in livers.

Motivated by these results, we then sought to examine if statins are similarly associated with a reduced risk of cancer development in human patients. To this end, we conducted a retrospective, two-center nested case-control study, wherein we evaluated all statin-taking patients relative to control groups at two large university hospitals—one in the U.S. (Virginia; UVA) and the other in Germany (Charité). Finally, we extended those RWE results by also utilizing a clinical research platform (TriNetX) to access a large, international cohort of anonymized EHR data (aggregate statistics only).

### Statins Are Associated with Reduced Cancer Risk

We considered an initial total of 308,990 patients with enrollment dates between 2008 and 2018. After data pre-processing (Figure 1), 277,980 patients were included in our RWE study. The trans-Atlantic cohort was first examined without further exclusion criteria, to test if statin intake might be linked to a cancer preventive effect. For this reason, a collection of statins was considered, thus increasing the group of cancer patients with statin intake; among these, simvastatin and atorvastatin were the two most commonly prescribed statins, accounting for 71% and 19% of the study populations, respectively. Other prescribed statins included pravastatin (5%), fluvastatin (2%), rosuvastatin (2%) and lovastatin (1%).

Most importantly, we found that statin intake correlates with a meaningful, statistically-significant reduction in the incidence of cancer (OR, 0.72; 95% CI, 0.70–0.74) in the trans-Atlantic cohort. For patients taking statins, we calculated a higher probability of surviving cancer versus subjects who were not taking any statins (HR, 0.64; 95% CI, 0.48–0.86). In considering if dosing levels influence the survival probabilities, we found no difference for low-dose (20mg: HR, 1.1; 95% CI, 0.89–1.37) versus high-dose (80mg: HR, 1.11; 95% CI, 0.83–1.48) treatments of statin. Similarly, treatment with atorvastatin at low doses (10-20mg: HR, 0.80; 95% CI, 0.59–1.09) did not differ substantially from a high-dose regiment (80mg: HR, 0.72; 95% CI, 0.51–1.02). Based on the large number of patient records that featured both a prescribed statins as well as a cancer diagnosis, each statin was further considered separately. The results show a strong cancer-preventive effect for atorvastatin (OR, 0.41; 95% CI, 0.38–0.43) and also significant effects for fluvastatin (OR, 0.7; 95% CI, 0.57–0.85), pravastatin (OR, 0.63; 95% CI, 0.56–0.71) and rosuvastatin (OR: 0.43, 95% CI, 0.36–0.51); in contrast, simvastatin showed only a weak cancer preventive effect (OR, 0.9; 95% CI, 0.87– 0.94). We were unable to readily assess any effects of lovastatin because of a large confidence interval and smaller number of patients with this statin (Supp Figure 2).

We also evaluated the clinical data by considering it as a 1:1 matched-study design, using propensity score–matched sub-cohorts; this was performed in order to better control for confounding or spurious associations that might stem from different distributions of age and gender between the whole dataset and the subset of cancer patients (Supp Figure 1). As shown in Figure 4, we discovered further evidence supporting the cancer preventive effect of statins: (i) all statins considered together had an OR of 0.5 (95% CI, 0.48–0.51), while (ii) atorvastatin exhibited an OR 0.3 (95% CI, 0.28–0.32), fluvastatin’s OR was 0.65 (95% CI, 0.47–0.88), lovastatin’s OR was 0.51 (95% CI, 0.38–0.7), pravastatin’s OR was 0.47 (95% CI, 0.42– 0.54), rosuvastatin’s OR was 0.32 (95% CI, 0.26–0.38), and simvastatin featured an OR of 0.63 (95% CI, 0.61–0.66).

**Figure 4:**
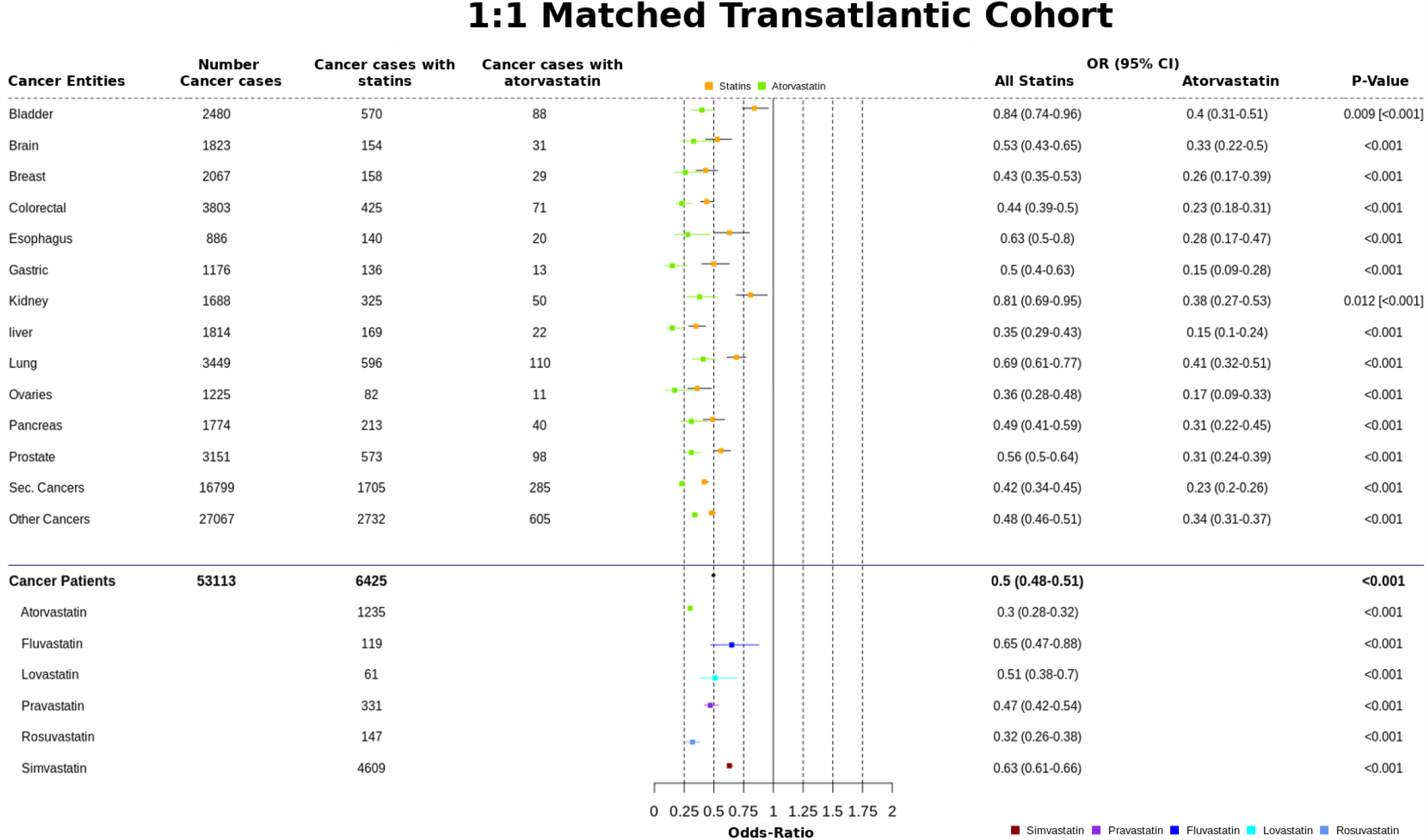
Cancer preventive effect of statins in the 1:1 matched trans-Atlantic cohort for different cancer entities, and comparison with different statins separately. The cancer preventive effects of statins as a group, and for atorvastatin separately, were calculated for a number of different cancer types, as indicated in this summary overview of (i) cancer incidences, (ii) cancer diagnoses and (iii) prescribed statins. Cancer preventive effects are calculated as odds-ratios (OR) for both statins and atorvastatin. The 95% CIs are provided in addition to the *p*-values; *p*-values are provided in square brackets for atorvastatin, for any values that differ from the full statin set. An overview for the different statins prescribed in the study population is provided at the bottom of the figure. Note the significant and systematic deviation—considered across different cancers—for atorvastatin in comparison to other statins.

### The Chemopreventive Effect of Statins Persists upon Exclusion of Confounders

To further account for potential confounders in our analysis, we also examined the drugs most commonly prescribed in conjunction with statins (Supp Figure 2), including calculation of OR values. Besides the statins, only aspirin and furosemide exhibit ORs < 1. All other co-medications do not show any cancer-preventive effect and were not further considered in this analysis. After excluding both drugs (aspirin and furosemide) in our 1:1 matched study design, we still found a cancer-preventive effect for all statins (OR of 0.756, 95% CI 0.72–0.80) as well as for atorvastatin alone (OR of 0.63; 95% CI, 0.57–0.70) (Supp Table 1). In a second step, the statin-specific co-diagnoses with cancer were calculated and sorted in descending order of appearance/occurrence (Supplementary Table 2). The overall outcome shows a slight increase of the odds ratio, but the cancer-preventive effect of statins remains unaffected. For some diagnoses, the OR values changed from approximately 0.5 to a maximum OR of 0.64 (95% CI, 0.61–0.68), that maximum being for hypercholesterolemia (ICD-10 codes E78 and Z83). From all of the above, we conclude that there is a real (statistically significant) signal indicative of chemopreventive effect for statins, despite any influence of confounders.

## Discussion

This study extends our previous work [5] by corroborating, broadening (via EHR data) and deepening (via MACC1-based experiments) our prior knowledge about the potential cancer-preventive effect of statins. On the molecular level, anti-cancer effects are likely mediated, at least partly, via inhibition of the tumor-promoting and metastasis-inducing gene *MACC1*. The MACC1 mRNA and protein is a known prognostic and predictive biomarker for 22 different solid-tumor entities, and it has been established as playing a key role in several processes that are signature events in cancer progression [22]. In this work, we found that most clinically employed statins diminish MACC1 expression.. In addition, we found a statin-induced reduction of MACC1 expression in CRC, as well as in pancreatic cancer and gastric cancer cells, thus confirming the cross-entity–wide effects in our RWE study. We also found that the chemopreventive influence of simvastatin (OR 0.63, 95% CI 0.61–0.66) and atorvastatin (OR 0.3, 95% CI 0.28–0.32) correlate with their capacity to reduce MACC1 expression, with IC_50_ values of 3.1 µM and 1.6 µM for simvastatin and atorvastatin, respectively (Supp Figure 5F).

Our *in vitro* experiments show that, in addition to cellular migration and wound healing (as reported in our previous study), MACC1–driven proliferation and colony formation are processes that are specifically inhibited by statins. More importantly, statin treatment led to a strong decrease of tumor growth, metastasis formation and MACC1 transcript levels *in vivo*, at dosages equivalent to a standard statin treatment regimen for blood lipid reduction [19,20]. In seeking to extend these molecular and experimental results to humans and to a broad population, we appealed to an RWE approach; in so doing, we used a strategy of a trans-Atlantic study design in order to ‘wash-out’ spurious factors (confounders) that might be responsible for differentially associating statins ↔ cancer within populations in the two environments (Germany, U.S.). The RWE revealed a significant link between statin usage and cancer development in patients. We found that statins exert cancer-preventive effects (with up to 50% cancer risk reductions), with specific patterns relating different cancer types and different statins. Analyses of individual cancer entities revealed a superior chemopreventive potential of statins for particular cancers, such as liver (OR 0.35, 95% CI 0.29– 0.43), colorectal (OR 0.44, 95% CI 0.39–0.50) or secondary neoplasms (OR 0.42, 95% CI 0.30–0.45).

As regards conflicting literature reports about statins in cancer therapy and prevention, this work offer (i) real-world evidence supporting the anti-cancer properties of statins, and (ii) experimental data that elucidates, in molecular terms, a potential role for the MACC1 protein in mediating a link between statins and at least some types of cancers. A recent analysis of 38 meta-analyses [11] obtained contradictory results: while some studies identified a preventive effect of statins for liver cancer (e.g., [24,25], with an OR of 0.58), others showed only a weak effect for CRC (OR 0.88 [26] and 0.94 [27]), or a conflicting trend for gastric cancer (ORs of 0.56 and 1.37, respectively, in [28,29]). Here, we report a strong risk-reduction for these other two (non-CRC) types: for liver cancer we find an OR of 0.35 (95% CI 0.29–0.43), and for gastric cancer the OR is 0.50 (95% CI 0.40–0.63). Another study found no increased risk to develop CRC [30] when taking statins, whereas other studies clearly demonstrated a reduced cancer risk for this entity [8,31-33]. These results are supported by a systematic review [34] and finally by our RWE data, with an OR of 0.44 (95% CI, 0.39–0.50). The recent New England Case Control study, which includes statin medication for at least six months, identified a reduced risk (OR 0.68) of ovarian cancer [6] and is corroborated by our RWE analysis, where we find an OR of 0.36 (95% CI, 0.28–0.48).

Because of our study design and large cohort size, we were able to scrutinize different statins separately: The statins with the highest case counts (simvastatin at 4,609 and atorvastatin at 1,235) exhibited differences in their risk-reduction potentials, with atorvastatin (OR 0.30, 95% CI 0.28–0.32) clearly exceeding simvastatin (OR 0.63, 95% CI 0.61–0.66). This might be of interest for future clinical trials or cancer therapy approaches, especially given that these are two of the most commonly prescribed statins [35,36]. Our study design also allowed us to exclude possible interfering factors, such as co-medication and co-diagnosis. Even aspirin co-medication, as the strongest confounder, only minimally influenced our results. Because we analyzed two completely separate and independent datasets (Charité, Berlin and UVA Hospital), as well as an international cohort (TriNetX), we mitigated the potential impacts of artifacts arising from regionally different prescription patterns.

As for any retrospective, RWE-based study, our observational analyses are not without limitations. For example, our results could be biased by undiagnosed cancers or by misclassification via the ICD10-catalog. We believe such effects are mitigated by ‘averaging out’ over our large cohort size and the inclusion of two independent cohorts, one in Germany and the other in the U.S. Nevertheless, an interesting future direction may be to utilize EHRs to try emulating a randomized target trial that would (theoretically) correspond to the observational analysis reported here, e.g. using Dickerman et al.’s recently developed framework [37]. Another limitation concerns longitudinal effects: because of a lack of patient follow-up data, we were limited to detecting potential cancer-preventive effects only based upon discharge data. Somewhat similarly, a lack of information on the duration of statin usage could influence the outcome in retrospective, observational clinical data such as those used in our RWE analysis. These caveats and limitations are mitigated by at least two important features of our study, namely that (i) we analyzed RWE from multiple independent streams (two trans-Atlantic cohorts and the multi-site TriNetX data-stores), and (ii) we examined the molecular basis for a potential mechanism of action of statins, wherein they act on MACC1 both in vivo and in vitro.

In conclusion, the RWE study reported here is consistent with prior work linking statins to cancer prevention. In addition to substantiating (and expanding) those earlier reports, we find that various confounding factors must be taken into consideration in order to cleanly distinguish artifacts from real effects. Based on our experimental results, we suggest a mechanism of action wherein statins act as chemopreventive agents by targeting MACC1 (or at least a MACC1-associated pathway); this model, in turn, highlights the significance of MACC1 as a decisive biomarker for patient stratification, and perhaps even as a molecular target for cancer therapy. How, specifically, might statins target MACC1? We have already shown, via electrophoretic mobility-shift assays and molecular docking analyses, that statins can inhibit MACC1 transcription by interrupting the binding (not the expression) of the essential transcription factors c-Jun and Sp1 to the MACC1 promoter, leading to reduced cell motility [5] and restored treatment response [38]. In addition, a further mechanistic answer may exist, albeit indirectly: A pattern of pathway linkages between statins and MACC1 can be found by recognizing known physiological connections between (i) fat/glycogen metabolism, which is well-established as being statin-related (e.g. the phenomenon of statin muscle myalgia), (ii) the known role of stress-response protein HSPA5 (heat shock 70kDa protein 5, also known as Grp78) in fat metabolism, (iii) a recently discovered (molecular) link between HSPA5/Grp78 expression levels and statin treatment [39], and (iv) a bioinformatic analysis [40] that suggested HSPA5 may be associated with SH3-domain binding protein 4 (SH3BP4). As the final piece of this puzzle, note that MACC1—which also harbors an SH3 domain—is the only paralog of SH3BP4 (MACC1 is cross-referenced in some databases as an “SH3BP4-like” protein), thus offering a chain of (secondary) linkages from the phenotypic level (i.e., statin use) to the molecular and pathway level of MACC1 itself. Though speculative, this model of statin ←…→ MACC1 linkages does offer mechanistic and testable hypotheses, and, perhaps eventually, ideas for new therapeutic directions.

To transform our findings into practical solutions for routine patient care, the known side-effects of statins, such as muscular syndromes (e.g., myalgia, myositis or rhabdomyolysis), would need to be evaluated on a case-by-case basis, ideally as part of the assessment regarding prescriptions and cancer prevention. However, the lack of validated diagnostic tools or clinical criteria, besides creatinine levels, impedes a precise prediction of (or even evaluation of) statin side-effects [41]. We hope the present study is useful in this regard, by providing RWE for the anti-cancer effects of statins, as well as in vitro and in vivo experiments that elucidate the molecular basis of these effects vis-à-vis the cancer-driving MACC1 protein. Before suggesting consideration of different statin treatment regimens (e.g., switching from simvastatin to atorvastatin) in order to broaden the scope of the chemopreventive effect of statins in the population at large, we suggest that targeted prospective trials may be beneficial, with patients potentially stratified based upon expression levels of the MACC1 biomarker.

## Data Availability

All data referred to in the manuscript is available either as supplementary materials or by request from the study's corresponding authors.

## Acknowledgements

We thank members of our laboratories for helpful discussions about this work.

