## Supplemental File for "Real-world Evidence for Preventive Effects of Statins on Cancer Incidence: A Trans-Atlantic Analysis"


This Supporting Information PDF file provides the following three sections of content:

- a **Supplementary Methods** section (provided herein)
- full captions for the **Supplementary Figures** (which are provided as separate image files in a Zip archive, with filename "`Gohlke_Statins_SuppInfo_raw-files_jun2021.zip`")
- synopses of the three **Supplementary Tables** (which are provided as data files in a Zip archive, with filename " `Gohlke_Statins_SuppInfo_raw-files_jun2021.zip`")

Please note: the Supp Figure image files and Supp Tables are provided as the auxiliary Zip archive (mentioned above).

**Supplementary Methods**: More detailed descriptions of our experimental and computational methods

### Cell Lines and Growth Conditions

Human cancer cell lines HCT116 (CRC), HT-29 (CRC), SW48 (CRC), BxPC3 (pancreatic cancer) were purchased from the American Type Culture Collection and MKN45 (gastric cancer) was kindly provided by Experimentelle Pharmakologie & Onkologie Berlin-Buch GmbH (EPO GmbH, Berlin, Germany). Cells were grown in DMEM or RPMI1640 medium (Thermo Fisher Scientific, Waltham, Massachusetts) supplemented with 10% fetal bovine serum (Bio & Sell, Feucht, Germany) in a humidified incubator at 37 °C with 5% $CO_2$. Cell lines were regularly tested for mycoplasma using the MycoAlert Mycoplasma detection kit (Lonza, Basel, Switzerland). We verified the authentication of all cell lines by short tandem repeat (STR) genotyping at the Leibniz-Institute DSMZ (Braunschweig, Germany); STR genotypes concurred with published genotypes.

Drug treatment was performed with statins purchased from either Selleckchem (Houston, Texas) (atorvastatin) or Thermo Fisher Scientific (Waltham, Massachusetts) (fluvastatin, simvastatin, lovastatin). The 20 mM stock solutions were prepared freshly, for every application, in dimethylsulfoxide (DMSO). Control cells were treated with equal amounts of solvent to rule out adverse effects caused by DMSO.

### Transfection of Human CRC Cells

Cell line generation for HTS was described previously[1,2]. For constitutive or inducible MACC1 overexpression, cells were transduced with lentiviruses produced in HEK293T cells using the plasmids RC224774L2 (MACC1-GFP, Origene, Rockville, Maryland) or pCW-GW-rtTA/MACC1-P2A-nLuc (pCW-GW-rtTA-P2A-nLuc was kindly provided by Dr. Nikolas Gunkel, DKFZ, Heidelberg) and pMD2.G, psPAX2 for packaging following standard protocols. pMD2.G and psPAX2 were kind gifts from Didier Trono (Addgene plasmid # 12259, # 12260). Cells were selected either by FACS or blasticidin (InvivoGen, San Diego, California) treatment. Knock-out of MACC1 in HCT116 cells (HCT116/MACC1-/-) was performed according to the protocol by Ran et al. [3] with the vector pSpCas9(BB)-2A-GFP (Addgene, Watertown, Massachusetts) and the single guide RNAs (sgRNAs) : sgMACC1 fwd – GTT TGA AGA GTA CCC GGG TTT GG and sgMACC1 rev – ACA TGC CTT GCT CCG TAT GCA GG (Biotez, Berlin, Germany) [3].

### High-throughput Drug Screening (HTS)

We seeded HCT116-MACC1p-Luc CRC cells stably expressing the MACC1-promoter driven luciferase gene in 384-well plates (Perkin Elmer, Waltham, Massachusetts) at 4,000 cells/well. The plates already contained the 4241 compounds in two different concentrations: 5 µg/ml and 0.5 µg/ml of Prestwick library compounds, and 1 µM and 0.1 µM of NIH and Microsource library compounds, respectively. Cells were incubated with compounds for 24 h at 37°C in a humidified incubator with 5% $CO_2$. To assess the luminescent signal, plates were treated with 25 µl BriteLite plus luminescent reagent for

5 min (Perkin Elmer, Waltham, Massachusetts) and measured in an Envision Reader with ultrasensitive luminescence detector (Perkin Elmer, Waltham, Massachusetts).

### In Vivo Validation of Statins

Animal experiments were conducted according to the United Kingdom Coordinating Committee of Cancer Research (UKCCCR) guidelines and granted by the State Office of Health and Social Affairs (Landesamt für Gesundheit und Soziales, LaGeSo, Berlin, Germany).

We transplanted $3x10^5$ of HCT116/CMVp-Luc cells into the spleens of 6-week-old female SCID-beige mice (Charles River, Wilmington, Massachusetts). Mice were randomly assigned to 3 groups of 10 animals for oral application with daily doses of either solvent (10% Kolliphor in 0.9% NaCl) or a human equivalent dose of approximately 1 mg/kg body weight (13 mg/kg body weight) of fluvastatin or atorvastatin (in 0.9% NaCl). This dose is widely used in blood lipid reduction therapy [4,5]. We continuously monitored tumor growth and metastasis formation via intraperitoneal application of 150 mg/kg D-luciferin (Biosynth, Staad, Switzerland) and the bioluminescence imaging system NightOWL LB 981 (Berthold Technologies, Bad Wildbad, Germany). For imaging and quantification, WinLight (Berthold Technologies) and ImageJ (version 1.51j8, National Institutes of Health, USA) were used. Afterwards, spleens and livers were removed and shock frozen in liquid nitrogen for preparation of cryosections and further analysis. Extraction of RNA with subsequent qRT-PCR analysis was performed as described in the next section.

### RNA Extraction and qRT-PCR

Drug treatment was performed in 12-well dishes with $1.25x10^5$ cells/well. To isolate total RNA, we used the Universal RNA Purification Kit (Roboklon, Berlin, Germany) according to manufacturer´s instructions. RNA was quantified (Nanodrop, Peqlab, Erlangen, Germany) and 50 ng RNA was applied to reverse transcription with random hexamers in a reaction mix (5 mM MgCl2, 1x RT-buffer, 4 mM pooled dNTPs, 1 U/µl RNAse inhibitor and 2.5 U/µL Moloney Murine Leukemia Virus reverse transcriptase; Thermo Fisher Scientific) at 23°C for 15 min, 42°C for 45 min, 99°C for 5 min with subsequent cooling at 4°C for 5 min. The LightCycler 480 (Roche Diagnostics, Mannheim, Germany) with GoTaq dye (Promega, Fitchburg, Wisconsin) chemistry was employed for cDNA amplification in a quantitative polymerase chain reaction (qPCR) under the following conditions: 95°C for 2 min followed by 45 cycles of 95°C for 7 s, 60°C for 10 s and 72°C for 5 s. Primers for MACC1 and G6PD were described previously [6]. Data were analyzed with the LightCycler 480 Software release 1.5.0SP3 (RocheDiagnostics). Duplicate qRT-PCR reaction values were averaged and each mean value of the expressed gene was normalized to the respective mean amount of G6PD cDNA.

### Protein Extraction and Western Blotting

Similar to RNA extraction, $1.25x10^5$ cells were seeded in 12-well plates. After drug treatment, cells were scraped off in ice cold RIPA buffer (50 mM Tris, 150 mM NaCl and 1% Nonidet P-40; pH 7.5 supplemented with complete protease inhibitor tablets; Roche Diagnostics) and lysed for 30 min on ice. We used the Bicinchoninic Acid Protein Assays Reagent (Thermo Fisher Scientific) to determine protein concentration according to manufacturer´s instructions. Equal amounts of protein were separated by SDS-PAGE and transferred to PVDF membranes (Bio-Rad Laboratories Inc., Hercules, California). Membranes were blocked for 1 h at room temperature with 5 % BSA in TBST buffer (10mM Tris-HCl, 0.1% Tween20 and 150 mM NaCl; pH 7.5). Incubation of membranes with rabbit anti-MACC1 antibody (Sigma-Aldrich, dilution 1:10,000) or mouse anti-β-actin (Sigma-Aldrich, dilution 1:25,000) and vinculin (Sigma-Aldrich, 1:2,000) at 4°C overnight was followed by incubation with HRP-conjugated anti-rabbit IgG (Promega, dilution 1:20,000) or anti-mouse IgG (Thermo Fisher Scientific, dilution 1:20,000) for 1 h at room temperature. Antibody-protein complexes were visualized with WesternBright ECL HRP substrate (Advansta, Menlo Park, California) and subsequent exposure to CL-Xposure Films (Thermo Fisher Scientific). Immunoblotting for β-actin and vinculin served as protein loading control.

**Proliferation Assay**

Cell viability and proliferation was assessed using the IncuCyte® ZOOM System (Essen BioScience, Ann Arbor, Michigan). $5x10^3$ cells were seeded in 96-well plates and allowed to accommodate for 24 h before drug treatment. After drug treatment, plates were placed into the IncuCyte for 72 h at 37°C in a humidified incubator with 5% $CO_2$. Four pictures of each well were taken every 2 h which allowed a detailed analysis of the area covered by proliferating cells over time. For data analysis and export the IncuCyte ZOOM software 2016B was used. Each cell proliferation experiment was performed in triplicate. Results are expressed as area under the curve of cell confluence normalized to solvent-treated controls.

**Clonogenic Assay**

We used the clonogenic assay to assess the reproductive viability and ability to form colonies. 400 cells/well of HCT116/GFP and HCT116/MACC1-GFP cells were seeded in 6-well dishes and allowed to attach for 24 h before drug treatment. After drug treatment, plates were placed into a humidified incubator at 37°C with 5% $CO_2$ for 7 days. Medium was removed and the colonies were fixed and stained with a solution of PBS containing 1% formaldehyde and 0.1% crystal violet. The colony covered area was determined with the colony area plug in and colony number with the particle analysis function of ImageJ (version 1.51j8, National Institutes of Health, USA). Each clonogenic assay was performed in triplicate. Results are presented as total counts or after normalization to solvent-treated controls.

### **References** (for Supp Methods)

---

### Supplementary Figure legends

#### Supplementary Figure 1: Age and gender distribution in the transatlantic cohort and patients diagnosed with cancer.

The age distribution for both genders in the full transatlantic cohort is shown (left half), along with the analogous distribution for patients diagnosed with cancer (right); any effects that might stem from the visible differences between these two groups were mitigated by using a 1:1 matched study design.

#### Supplementary Figure 2: Cancer preventive effect of different statins and different cancers across the transatlantic cohort.

The cancer preventive effect of statins as a group, and atorvastatin alone, were calculated for a number of different cancer types. A detailed overview of cancer incidences, cancer diagnoses and prescribed statins is provided. The cancer preventive effect is calculated as an odds-ratios for both, statins and atorvastatin. The 95% confidence intervals p-values are provided. P-values in square brackets are provided for atorvastatin, for any values that differ from the statins at large. An overview of results for all statins prescribed in the study population is presented in the bottom of the figure.

#### Supplementary Figure 3: Co-medication with statins

The medications prescribed alongside statins are shown, in decreasing order of the total number of prescriptions of each.

**Supplementary Figure 4: Statins reduce MACC1 expression in different cancer entities.**

Fluvastatin and atorvastatin decreased MACC1 mRNA and protein expression in pancreatic cancer (BxPC3; Panel A) and gastric cancer (MKN45; Panel B) cells. MACC1 mRNA levels were normalized to G6PD mRNA expression and respective treatment controls (DMSO, indicated with white bars). Results for mRNA represent means + Standard error of the mean (SEM) of three independent experiments and for WB one representative example of at least two independent experiments is shown. In the WB, β-actin or vinculin served as loading control. Significant results were determined by one-way ANOVA and Dunnett's multiple comparison test with a confidence interval of 95% (* = $p < 0.05$, ** = $p < 0.01$, *** = $p < 0.001$, **** = $p < 0.0001$).

**Supplementary Figure 5: Statins specifically inhibit MACC1-mediated functions in vitro.** Relative proliferation was determined with the IncuCyte® live imaging system for 72 h and calculated by the area under the curce (AUC) normalized to untreated controls (white bars). HCT116 cells with doxycycline induced MACC1 expression (+Dox) demonstrated a 30% increase in proliferation. MACC1-induced proliferation in +Dox compared to -Dox cells was still observed under statin treatment indicating a MACC1-specific rescue of proliferative function (Panel A). The previous findings were confirmed using HCT116/MACC1 -/-. MACC1 knock-out resulted in an over 50% reduction in proliferation compared to control cells (HCT116/Cas9). Control cell proliferation was decreased by fluvastatin treatment (1 and 2.5 µM) whereas HCT116/MACC1 -/- cells remained unaffected (Panel B). Stable overexpression of MACC1-GFP in HCT116 led to a strongly augmented colony formation compared to HCT116/GFP cells. The same effect was seen under statin treatment indicating a MACC1-specific rescue of clonogenic function (Panel C). Clonogenicity was quantified by the number of colonies (Panel D) and colony covered area (Panel E). Results represent means + SEM of at least three independent experiments normalized to solvent treated controls (white bars) or presented as total counts (Panel D). For the clonogenic assay one representative of nine technical replicates of three independent experiments is shown. Significant results were determined by one-way or two-way ANOVA and Sidak´s multiple comparison test with a confidence interval of 95% (* = $p < 0.05$, ** = $p < 0.01$, *** = $p < 0.001$, **** = $p < 0.0001$). Panel F shows the relative MACC1 mRNA expression determined in Figure 3 presented as drug-response curves for IC50 determination. Fluvastatin (IC50: 0.8457 µM) is able to reduce MACC1 mRNA expression at lower doses as atorvastatin (IC50: 1.647 µM) and simvastatin (IC50: 3.098 µM).

**Supplementary Figure 6: Survey of the top-50 most-prescribed drugs in Germany in 2017**

Simvastatin can be seen to occupy position 16, and atorvastatin is at position 21 of the most prescribed drugs in germany. The data was collected by WIdO (PharMaAnalyst) and visualized by the Statista webportal. Data was accessed at the 15 of May 2019 (https://de.statista.com/statistik/daten/studie/787888/umfrage/verordnungsstaerkste-arzneimittel-in-deutschland/.).

**Supplementary Tables**: Synopses of the files (Excel files in Zip format):

- **Supplementary Table 1.xls**
  This table consists of the raw-results from the RWE study. Odds-Ratios and Relative-Risks are calculated for different statins and different cancer entities. Also, both cohorts--Charité and UVA--were considered together in the trans-Atlantic cohort as well as separately.

- **Supplementary Table 2.xls**
  This spreadsheet supplies the results of our analyses of co-medications prescribed with statins.
  The first tab “Co-medication Counts” provides a list of drugs prescribed together with statins. The total number of counts of those prescriptions is provided (in descending order) for the top 10 co-medications.
  The second tab “Co-medication General” consists of the results (odds-ratios [ORs]) for those 10 drugs provided in tab 1. The ORs are calculated for the transatlantic cohort as well as the 1:1 matched cohort design study. Besides the ORs, p-values are also provided. Drugs with an OR < 1 are highlighted in yellow.
  The third tab “Exclusive Drug Calculation” consists of the analysis of previously identified drugs (tab 2) with an OR < 1. Those co-medications were analyzed separately by excluding patients taking at least two drugs identified in tab 2.
  These analyses were performed for the transatlantic as well as the 1:1 matched cohort. In addition, we analyzed the two most prominent statins (simvastatin and atorvastatin) separately to point out differences between statins.
  As can be seen in the tables, atorvastatin outperformed simvastatin with respect to the ORs in the transatlantic as well as 1:1 matched cohort.

- **Supplementary Table 3.xls**
  In this file, the co-diagnoses of cancer were analyzed, taking into consideration statin intake as well.
  In tab 1 “Counts”, co-diagnoses are calculated. The resulting diagnoses are presented via a short description as well as the ICD10 code. “Counts” consists of the number of patients, also diagnosed together with cancer in descending order.
  The second tab “Odds-Ratios”, shows the ORs. The first results row displays the OR for cancer given (statins, atorvastatin, simvastatin) without exclusion of any co-diagnoses.
  The rows 5-11 and 16-22 shows the cancer preventive effect of statins by excluding the diagnosis in column B.
  Results are displayed for the transatlantic as well as 1:1 matched cohort.
